# Assessing ageing, cognitive ability and freezing of gait in Parkinson’s disease through integrated brain–heart network dynamics

**DOI:** 10.64898/2026.04.22.26351482

**Authors:** Laura Pitti, Giovanni Sitti, Diego Candia-Rivera

## Abstract

Parkinson’s Disease (PD) is a complex neurodegenerative disorder that manifests through systemic, large-scale physiological reorganizations. While research often focuses on region-specific neural changes, there is a growing need for multidomain approaches capable of capturing the complexity of the disease and its clinical heterogeneity. Here, we propose an analytical pipeline to evaluate Brain–heart Interactions (BHI) as a systemic biomarker of neurodegeneration and healthy ageing.

In this study, we assessed BHI across three open-source datasets combining EEG and ECG recordings. We compared Healthy Young adults, Healthy Elderly participants, and early-stage PD patients during resting state to investigate the effects of ageing and neurodegeneration on large-scale physiological organization and cognition. In addition, we examined BHI dynamics surrounding episodes of freezing of gait (FOG) in PD patients. Methodologically, brain network organization was quantified using coherence-based EEG functional connectivity and graph-theoretical metrics, while cardiac dynamics were characterized through Poincaré plot-derived measures of autonomic activity. Coupling between the two systems was estimated using the Maximal Information Coefficient, enabling the detection of both linear and non-linear dependencies between cortical network organization and cardiac autonomic outflow.

Our results demonstrate that brain–heart networks are sensitive to systemic physiological changes associated with both healthy ageing and early PD. Specifically, we observed distinct BHI profiles differentiating Healthy Young, Healthy Elderly, and PD participants, suggesting that the proposed framework captures progressive alterations in integrated neurophysiological regulation. Importantly, resting-state BHI metrics were associated with cognitive performance in early PD, supporting the relevance of brain–heart coupling as a marker of clinical heterogeneity beyond motor symptoms alone. Furthermore, the analysis of FOG episodes revealed the emergence of specific BHI interaction clusters preceding and during gait freezing, highlighting coordinated alterations in cortical and autonomic dynamics linked to motor dysfunction. Together, these findings suggest that brain–heart networks provide a promising systems-level framework for understanding PD symptomatology and detecting early multisystem dysfunction in neurodegeneration and ageing. Our proposed pipeline offers a scalable and clinically relevant tool for large-scale physiological assessment in translational and clinical neuroscience research.

**Graphical abstract:** Analytical pipeline to evaluate Brain–heart Interaction (BHI)
Simultaneous electroencephalogram (EEG) and electrocardiogram (ECG) recordings undergo advanced signal preprocessing, followed by the extraction of brain and cardiac autonomic indices. For the EEG, graph-theory-based network organization metrics are computed, including Global Efficiency (E_g_) and Modularity (Q). For the ECG, Poincaré analysis of inter-beat interval (IBI) sequences is used to derive cardiac sympathetic (CSI) and parasympathetic (CPI) indices. Coupling between EEG and ECG time series is then quantified using the Maximal Information Coefficient (MIC). This multimodal framework provides informative markers related to ageing, cognitive ability, and freezing of gait in patients with Parkinson’s disease.

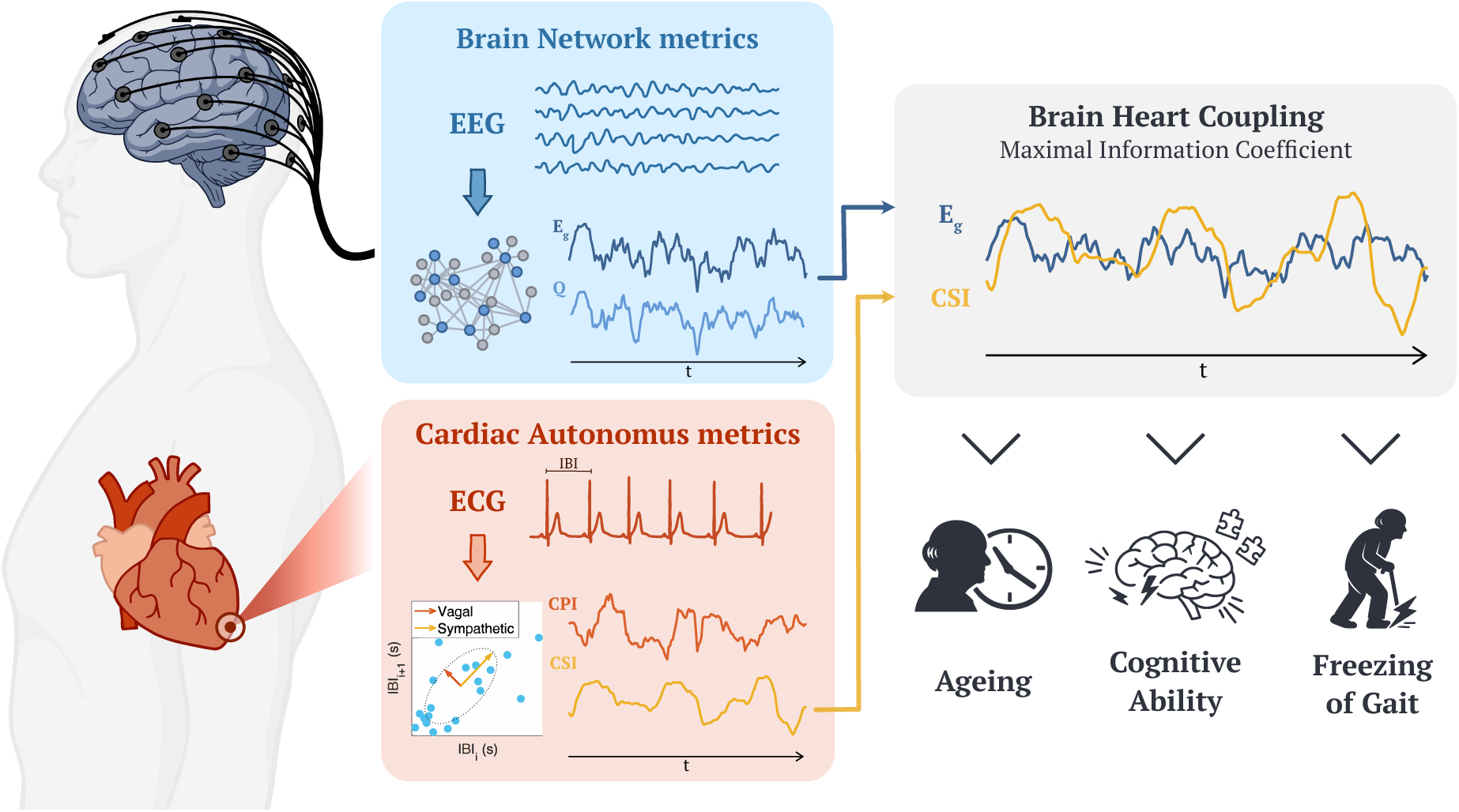

**Highlights:**

- We propose a pipeline based on EEG-ECG to assess ageing and neurodegeneration
- Brain–heart networks detect systemic changes in ageing and early PD
- Resting brain–heart networks relate to cognitive performance in early PD
- Specific brain–heart interaction clusters emerge during freezing of gait
- Brain–heart networks offer a promising tool to understand PD’s symptomatology

## Introduction

Parkinson’s Disease (PD) is characterized by a combination of motor and non-motor symptoms. Although the pathology is usually diagnosed upon the onset of the first motor signs, non-motor symptoms may manifest a decade or more before the clinical diagnosis [1]. These non-motor symptoms encompass a wide range of physiological alterations, including sleep disturbances, constipation, sensory loss (such as anosmia and ageusia), cognitive impairment and psychiatric symptoms, ranging from apathy to depression and anxiety [1], [2], [3]. Motor symptoms includes bradykinesia and hypokinesia, rigidity, tremor and postural symptoms as gait and balance disorders [2], [4], [5]. Freezing of Gait (FOG) is a highly disabling symptom defined as a brief episodes of inability to step or by extremely short steps that typically occur on initiating gait or on turning while walking [6]. These episodes can be caused by different triggers including turning, multi-task actions and emotional states.

PD is neuropathologically defined by loss of dopaminergic neurons and the presence of Lewy bodies and Lewy neurites in the substantia nigra, a brain region essential for the regulation of movement [1]. Long, thin, poorly myelinated axons with extensive branches, as the vagus nerve, are more vulnerable to degeneration. Indeed, studies have consistently reported that a high proportion of patients develop impairments within the autonomic nervous system, a condition commonly referred to as dysautonomia [3], [7], [8], [9]. Dysautonomia results in a large-scale and multisystem disruptions causing common symptoms like abnormal heart rate variability (HRV), reduction in orthostatic hypotension along with hypertension, dyspnea, gastroparesis, and incontinence [2], [3], [7], [8], [9], but it remains mostly unclear if and how these precedes and influence the motor and non-motor symptoms of the disease, included the FOG episodes [3], [10], [11], [12], [13]. This condition makes PD an ideal model for studying biomarkers for multidomain levels of characterization. While extensive research has demonstrated that autonomic dysfunction significantly contributes to the multimodal dysregulation observed in PD [14], research typically focus on their cognitive and motor symptoms. These autonomic impairments can be quantitatively assessed through HRV analysis or monitoring of blood pressure dynamics, which evaluate the physiological responsiveness of the autonomic nervous system to psychological stimuli. For instance, HRV analysis via ambulatory ECG showed potential for assessing autonomic dysfunction in PD patients [15], in which significant reductions in HRV appeared among untreated PD patients and correlated with disease severity.

The autonomic nervous system plays a crucial role in physiology and pathophysiology, serving as the fundamental link between the brain and peripheral organs. However, even in the absence of autonomic dysfunction, the role of cardiac autonomic dynamics in neurodegeneration remains to be understood. Recent research has shown that cardiac dynamics can influence neural states through continuous bidirectional interactions [16], [17], [18]. This suggests that PD triggers large-scale physiological reorganizations at the very early stages of the disease, preceding the manifestation of classic autonomic symptoms. PD patients exhibit highly heterogeneous symptoms and variable involvement of different neuronal systems during the early disease phases, making the development of a unified diagnostic or therapeutic approach particularly challenging [3], [19], [20]. This heterogeneity confirms PD as a network pathology, supported by existing evidence of alterations in brain connectivity and networks [4]. Consequently, research should move beyond region-specific analyses; adopting a large-scale perspective is essential to identify reliable biomarkers for such complex conditions.

Recent research in PD showed that quantifying the coupling between ongoing brain network organization and cardiac sympathetic–vagal oscillations provides an accurate estimator of the effectiveness of dopaminergic replacement therapy to reduce motor symptoms, in the absence of autonomic dysfunction [21], [22]. The present work leverages on this framework for the quantification of brain–heart interactions [22], where brain networks are characterized through coherence-based EEG connectivity and graph theory to derive metrics of global brain network organization. In parallel, cardiac dynamics are assessed via Poincaré plot geometry, a validated method for estimating time-varying cardiac sympathetic and parasympathetic activities [23]. The core of this investigation lies in quantifying the coupling between global brain network dynamics and cardiac fluctuations to capture complex linear and non-linear dependencies. This pipeline is aimed to provide comprehensive physiological insights into the complex interplay between brain and heart in normal ageing, cognitive performance and the motor complications of PD. This framework facilitates more consistent large-scale assessments and has the potential for the early detection of multisystem dysfunctions.

## Materials and methods

### Participants

Three open-source datasets have been used, two of them have been analyzed to study differences in age and cognitive ability, while the third was used to study the FOG.

The first dataset is from a previously published set from George et al. [2] downloaded from OpenNeuro [24] and previously studied by Appelhoff et al. [25], Jackson et al. [26], Pernet et al. [27], Swann et al. [28]. The dataset is composed by 15 PD patients, 8 female, with a mean age of 62.6 years, and 16 Healthy Elderly (HE) controls, 9 female, with a mean age of 63.5 years. Controls were chosen to match with PD in age, gender, handedness, and Mini-Mental Status Exam cognitive scores. For all Parkinson’s disease participants, data selection for the analysis was restricted to the off-dopamine medication condition. Specifically, the first dataset was acquired according to an Institutional Review Board Protocol at the University of California, San Diego [2].

The second dataset is from the study of Koelstra et al. [29] with 32 Healthy Young (HY) patients, 16 female, mean age of 26.9 years, recorded in resting state. All participants provided written informed consent prior to data acquisition.

The third analyzed dataset has been downloaded from Mendeley Data [30] and comes from the study conducted by Zhang et al. [31], with 12 participants, 6 female, mean age 69.1 years, with PD and FOG. Data collection was performed while patients were in the off-dopamine medication condition. This research has obtained the Ethical approval (N. 2019-014) from the Ethics Committee of Xuanwu Hospital, Capital Medical University, Beijing, China. The study has been conducted in conformity to Declaration of Helsinki and before the data recording written informed consents from all participants were obtained.

### Experimental setup

In both the first [2] and second [29] dataset Electroencephalogram (EEG) and Electrocardiography (ECG) were acquired. EEG data were recorded according to the international 10-20 system using a 32 channel ActiveTwo Biosemi system at a sampling frequency of 512 Hz. The analysis for this study was restricted to 2-minute resting-state sessions. During recording, subjects were instructed to relax and keep their eyes open, focusing on a central white fixation cross displayed on the monitor [2], [29].

In the third dataset [31] a multimodal sensory platform is used to acquire EEG, Electromyography (EMG), ECG and accelerometers (ACC). A 32-channel EEG was positioned following the international 10-20 system, along with EMG and ECG were acquired using a wireless MOVE system (MOVE, Brain Products GmbH, Gilching, Germany) with a sampling rate of 1000 Hz. ACC data were collected by MPU6050 6-DoF accelerometer and gyro and LM324. Four inertial sensors were mounted at the lateral tibia of the left and right legs, fifth lumbar spine (L5) of the waist and left arm, respectively. ACCs were sampled at the frequency of 500 Hz. To standardize the sampling frequency across all sensors, EEG was down-sampled to 500 Hz. Patients had to complete two different walking tests designed to induce freezing of gait, each test was repeated twice. The entire experiment was also recorded as a video to let two qualified physicians, from the Department of Neurology of Beijing Xuanwu Hospital, to label the start and end points of each FOG episode [31].

### Subject selection

Prior to the preprocessing and the analysis of signals there was the need to make a selection of the subjects in the dataset of PD patients with FOG [31]. The aim was to find events that were long enough and did not have any preceding FOG event in a defined time interval prior to the onset. Ahead of defining a selection threshold, FOG array has been cleaned relying on the following steps: first, if the interval between two consecutive FOG was lower or equal to 3s, then the two events were merged together; second, if the duration of FOG was less than 3s and the distance between that and the previous one was greater than 3s, then that FOG event was not considered.

For the remaining events, the selection thresholds were based on those established in a previous study [32]. We considered FOG events with a minimum duration of 15 seconds, preceded by at least 40 seconds of FOG-free activity.

The last step was to verify the integrity of the ECG signal within FOG events, ensuring that the ECG wave complexes remained clearly discernible and free of noise.

It has been defined a specific analysis window of 20 seconds where the onset, the time zero of the window, was shifted 5 seconds prior to the clinical FOG initiation [32]. Given this definition all the metrics about brain networks, cardiac autonomic dynamics and their interaction were calculated for both the pre-FOG interval, going from −10s to 0s, and FOG interval, from 0s to 10s.

### Preprocessing

EEG data were preprocessed in MATLAB-R2024a using the FieldTrip toolbox [33]. The preprocessing steps included a fourth-order Butterworth bandpass filter [1–45] Hz, a notch filter [49–51] Hz to remove power line interference, eye movements and cardiac artifacts removal using ICA and re-reference to a common average [34].

We implemented an ad-hoc pipeline for FOG to treat movement-related artifacts. During the ICA-based artifact rejection step, we also cleaned the data from gait and FOG artifacts [35], [36]. Very large artifacts were filtered at the independent components space using wavelet filtering. In addition, components showing significant correlations with accelerometers attached to the limbs were identified through correlation analysis and removed from the data. Bad channel removal was performed by visual inspection estimating channel correlation respect to each neighbor using Fieldtrip. Channels exhibiting a neighbourhood correlation lower than 0.6 were rejected. Then, EEG series were reconstructed, and remaining channels were interpolated using the triangulation method.

ECGs were processed using R-DECO toolbox [37]. R-DECO is an envelope-based algorithm built upon the Pan–Tompkins method [38]. It enables automatic R-peak detection by flattening the ECG signal and enhancing QRS complexes, thereby improving robustness against noise [37]. To ensure data integrity, this step was followed by a visual inspection to identify misdetections, with manual corrections applied where necessary. The resulting R-to-R interval durations were used to construct the Interbeat Interval (IBI) series for the subsequent characterization of autonomic dynamics.

### Metrics

#### Cognitive ability

To quantify the level of cognitive ability, participants completed the Mini-Mental Status Exam (MMSE). MMSE is an administered psychometric screening assessment of cognitive functioning commonly used to quantitatively assess cognitive impairment. It consists of a variety of questions which final score spans from 0 to 30 points. Three levels classify the cognitive impairment: 24-30 = no cognitive impairment; 18-23 = mild cognitive impairment; and 0-17 = severe cognitive impairment [39]. To date, however, researchers have highlighted that a cut-off score of 24 may not yield an adequate balance between sensitivity and specificity, particularly in individuals with at least 16 years of education [40].

#### Brain network characterization

Spectral and cross-spectral densities were calculated via Short-Time Fourier Transform using 2s Hanning-tapered windows with a 50% overlap. This approach provided a fluctuating characterization with a 1s temporal resolution. The coherence *COH*_*i,j*_[*f*] between two EEG time series, *x*_*i*_(*t*) and *x*_*j*_(*t*), is derived from their complex Fourier transforms, *x*_*i*_[*f*] and *x*_*j*_[*f*], as defined below:

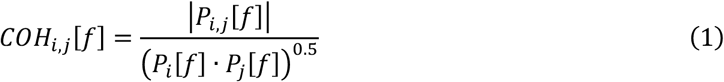

where *P*_*i,j*_[*f*] is the cross-spectrum of *x*_*i*_[*f*] and *x*_*j*_[*f*] and *P*_*n*={*i,j*}_[*f*] is the power spectral density of *x*_*n*={*i,j*}_(*t*). Functional connectivity was evaluated by integrating coherence within the alpha [8–12] Hz, beta [12–30] Hz, and gamma [30–45] Hz ranges. The resulting matrices were then thresholded using the efficiency–cost optimization algorithm to generate undirected binary graphs [22], [41]. Once undirected binary matrices were obtained for each time instant, global network metrics were calculated at each timestamp to achieve a time-varying estimation of network organization.

Global network dynamics were evaluated to characterize the intrinsic topological attributes of connectivity patterns from a region-agnostic, global perspective. In each connectivity matrix, rows and columns denote the nodes, where each voxel indicate the strength of the link between them [22], [42], [43], [44]. We focused on Global Efficiency (*E_g_*) and Modularity (*Q*) [22] using the Brain Connectivity Toolbox [44].

Global Efficiency typically indexes functional integration, as it captures how efficiently information is shared across the entire network architecture. Mathematically, is proportional to the inverse of the characteristic path length, the average minimum number of edges between any two nodes. Therefore it assumes that networks with fewer or longer connections exhibit lower communication efficiency [22], [43], [45].

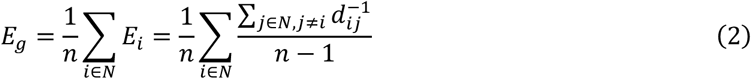

where *n* is the number of nodes in the network and *d*_*ij*_ is the distance between the nodes *i* and *j* quantified by the shortest path.

Modularity is a measure of network segregation that quantifies the degree to which a system is subdivided into distinct, non-overlapping communities or modules. A network with high modularity exhibits dense intra-module connectivity, where nodes within the same group are strongly linked, contrasted by sparse inter-module connections [22], [44]. Modularity was computed as follows:

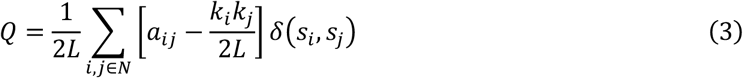

where *L* is the sum of all connectivity values, *a*_*ij*_ is a connectivity value between the nodes *i* and *j*, *k*_*i*_ is the degree of node *i*, and si indicates the group to which node *i* belongs. The term *δ*(*s*_*i*_, *s*_*j*_) is the Kronecker delta function, which is equal to 1 if *s*_*i*_ = *s*_*j*_, and 0 otherwise.

#### Alpha-gamma range

The selection of the frequency range spanning from the alpha to the gamma bands is primarily motivated by their strong correlation with resting state activity and higher cognitive functions. These bands are found to be sensitive to pathological changes associated with Mild Cognitive Impairment and aging [46]. Differences between healthy controls and patients are frequently detected in the alpha band during memory tasks [46], but also significant alterations in network organization, specifically regarding the balance between segregation and integration, have been documented in the beta and gamma bands [43], [47].

Our research focuses on the beta band for the study of Freezing of Gait (FOG). Beta-band oscillations are a physiological hallmark of normal motor circuits, a process that is significantly altered in patients with PD. Abnormalities include a failure in desynchronization, which is instead replaced by an hyper synchronization of specific brain regions [6]. long-duration temporal fluctuations of the beta rhythm, known as “beta bursts”, have been identified in the subthalamic nucleus and are particularly characteristic of ‘freezer’ patients [48]. Research also indicates an increase in local network segregation within the beta band across key areas, such as the frontoparietal cortex and the insula, during the transition to freezing [49]. This cumulative activity results in a system overload that manifests as a motor block. Consequently, this beta band oscillatory activity is considered the primary biomarker of impaired motor control in PD [10], [47], [48].

#### Cardiac Sympathetic and Parasympathetic indices

Cardiac sympathetic and parasympathetic activities were estimated over time using a method based on Poincaré plot geometry [26]. This method allows for a time-resolved estimation of autonomic oscillations through the calculation of the Cardiac Sympathetic Index (CSI) and Cardiac Parasympathetic Index (CPI). Such an approach facilitates a comprehensive exploration of dynamic shifts in autonomic regulation and their potential coupling with concurrent brain activity. It is demonstrated that while traditional spectral LF/HF measures provide a general overview, CSI and CPI offer a smoother and more robust estimator.

The Poincaré plot is constructed by plotting consecutive interbeat intervals (*IBI*_*i*_ versus *IBI*_*i*+1_) within a time window 𝛺_*t*_ of a defined duration *T*. These plots typically exhibit an ellipsoid-shaped distribution, characterized by three primary quantities that reflect specific physiological attributes of Heart Rate (HR) and HRV. The distance from the center of the ellipse to the origin (*CCD*) represents the baseline cardiac cycle duration. Meanwhile, the minor and major axes of the ellipsoid (*SD*_1_ and *SD*_2_, respectively) quantify the short- and long-term fluctuations of HRV. Therefore, the minor axis *SD*_1_ will capture high frequency variations of the heart rate, while the major axis *SD*_2_ reflects low frequency variations of HRV. The application of a sliding time window transforms static Poincaré parameters into dynamic indices. A window duration of *T* = 15 *s* provides an optimal balance, ensuring high temporal resolution while remaining capable of capturing the gradual fluctuations in HRV [50]. These parameters are computed as follows:

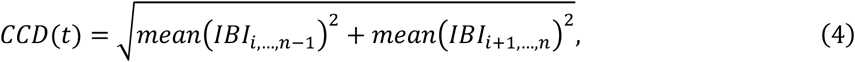

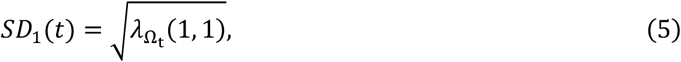

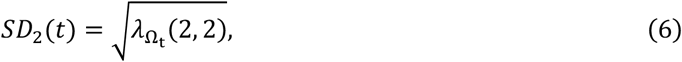

where 𝛺_*t*_: *t* − *T* ≤ *t*_*i*_ ≤ *t* is the sliding time window of duration *T*, *i* represent a generic first index of *IBI* in the time window and *n* is the length of *IBI* in the time window 𝛺_*t*_. Lastly, 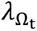 is the matrix with the eigenvalues of the covariance matrix of *IBI*_*i*,…,*n*–1_ and *IBI*_*i*+1,…,*n*_.

The CSI and CPI are derived from the integration of very slow heart rate dynamics and the respective slow and fast variability components. Specifically, CSI and CPI account for both the HR effect (represented by *CSI*_*HR*_ and *CPI*_*HR*_) and the specific contributions of HRV (*CSI*_*HRV*_ and *CPI*_*HRV*_). The indices are computed as follows:

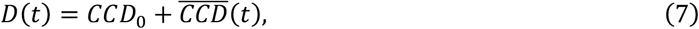

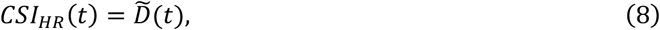

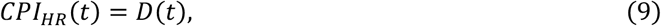

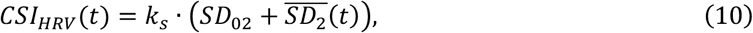

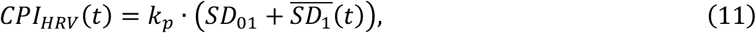

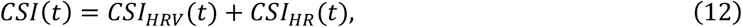

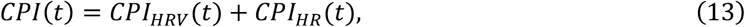

where the distance to the origin *CCD*_0_ and ellipse ratios *SD*_01_ and *SD*_02_ correspond to the computation on the whole recording, computed to re-centre the estimations of *CCD*, *SD*_1_ and *SD*_2_. The *SD*_0*x*_ is 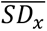 is the demeaned *SD*_*x*_ and *D̅* is the flipped *D* with respect to the mean. The coefficients *k*_*p*_ = 10 and *k*_*s*_ = 1 define the weight of the fast and slow HRV oscillations, with respect to the changes in the baseline *CCD*.

#### Maximal information coefficient

In the context of physiological coupling and non-linear dynamics, the most principled approaches are those framed within the field of information dynamics [51]. To quantify the interplay between brain network metrics and cardiac dynamics, we employed the Maximal Information Coefficient (MIC), a robust non-parametric statistic belonging to the Maximal Information Nonparametric Exploration (MINE) family [22], [51], [52].

MIC is designed to capture both linear and non-linear associations by identifying the maximum mutual information achievable through an optimal grid partition of the scatterplot. The fundamental principle of MIC is that if a relationship exists between two variables, a grid can be drawn on their scatterplot to encapsulate the data distribution. Formally, for a pair of time series X and Y, the algorithm explores all x-by-y grids up to a maximal resolution *B*(*N*), where *N* is the sample size. For each grid, the mutual information *I*_g_ is computed based on the probability distribution induced by the data points falling within the grid boxes. These values are then normalized by the minimum joint entropy log_2_ min {*n*_*x*_, *n*_*y*_} resulting in an index in the range 0 − 1 that ensure comparability across different grid dimensions. The characteristic matrix is constructed:

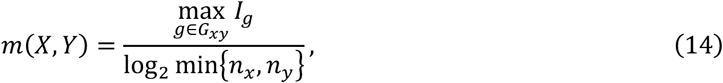

The MIC value is defined as the maximum entry in this matrix:

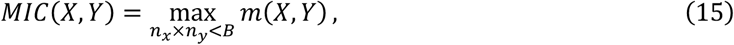

where the maximal resolution *B* is function of sample size *N* and is set following the standard literature recommendation, *B* = *N*^0.A^.

A primary advantage of MIC is that it does not require symbolic transformations of the data. Furthermore, a key advantage of MIC is its equitability; unlike the Pearson correlation coefficient, MIC assigns similar scores to relationships with similar noise levels regardless of the functional form (e.g., sinusoidal, linear, or parabolic) [52].

While brain–heart interaction is quantified using the MIC, which yields a single value per subject (ranging from 0 to 1), cardiac sympathetic–parasympathetic dynamics and brain network metrics are computed as time series. To obtain a representative value for each participant, these metrics were averaged across the entire recording duration.

### Statistical analysis

Non-parametric tests were used for the statistical analysis. Age-related differences between groups and correlations with age and cognitive ability were evaluated. Given the small sample size of the FOG dataset, we performed qualitative analyzes over the computed metrics.

Age related differences in brain, heart and brain–heart interaction among the groups were statistically evaluated using Kruskal-Wallis for multi-group comparison, and Wilcoxon rank-sum test for pairwise, unpaired comparisons: (i) healthy young and healthy elderly; (ii) healthy young and PD; and (iii) healthy elderly and PD participants. The Kruskal-Wallis test was employed to assess overall differences across the three groups (healthy young, healthy elderly and PD). Relationships between MMSE score and age with brain, heart and brain–heart interaction were evaluated using Spearman correlation coefficients.

The initial level of significance was set at *α* = 0.05. To ensure the reliability of the correlations and rule out spurious associations, a non-parametric Monte Carlo permutation test was implemented with number of iterations *N*_*iter*_= 10,000 [53]. The final significance was determined by calculating the Monte Carlo p-value (*p*_*mc*_), defined as proportional to the number of permutations for which the associated p-value was better (i.e. lower) than the original sequence (*N*_*pbest*_).

## Results

### Ageing

Brain network organization, cardiac autonomic dynamics and brain–heart interaction were evaluated in the healthy young, healthy elderly and PD groups. Results from the Kruskal-Wallis test are reported in Table 1 (see Supplementary Material, Table S1 for details on the most relevant brain network differences obtained via Wilcoxon rank-sum tests). Specifically, significant group differences were observed in brain–heart interaction metrics, particularly regarding the coupling between beta efficiency-sympathetic activity (*p* = 0.0032, *χ*^2^ = 11.0034) and gamma efficiency-sympathetic activity (*p* = 0.0003, *χ*^2^ = 13.9019). Although statistical significance was maintained when considering cortical and cardiac components independently, their discriminative power varied. Brain alone has demonstrated to have the highest discrimination capacity, but brain–heart interaction metrics exhibited a superior capacity compared to cardiac indices alone, as illustrated in the statistical comparison in Figure 1.

**Figure 1.**
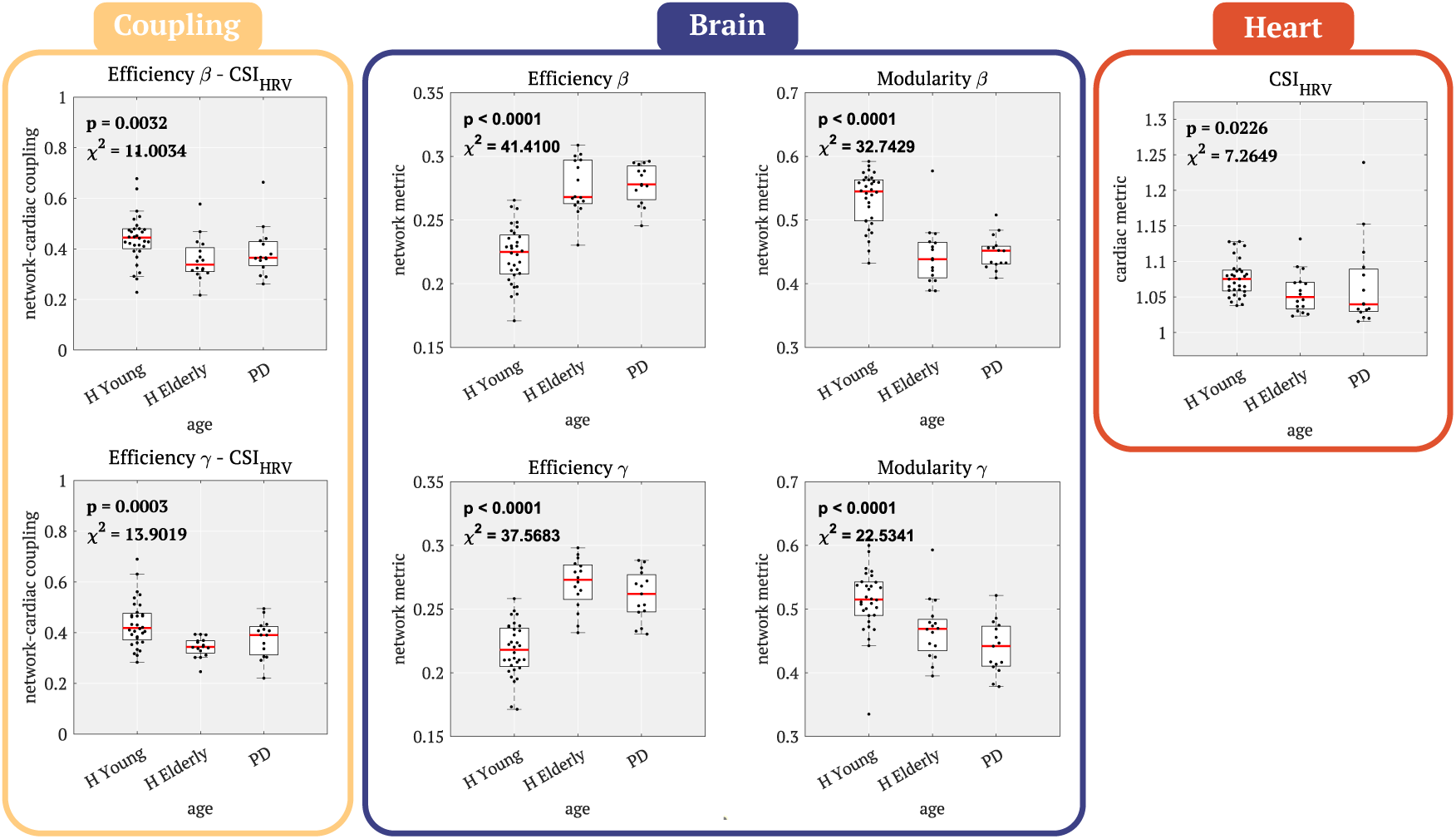
Statistical comparison of cortical, cardiac, and brain–heart interaction metrics. Group differences (HY, HE, and PD) evaluated via Kruskal-Wallis test. Yellow Panel (Coupling): Brain–heart Interplay indices for β-Efficiency-CSI_HRV_ and γ-Efficiency-CSI_HRV_. Blue Panel (Brain): Independent cortical metrics for β and γ bands, comparing integration (Efficiency) and segregation (Modularity). Red Panel (Heart): Isolated cardiac sympathetic variability index showing a significant group effect.

**Table 1.** Statistical results of Kruskal-Wallis test on differences between healthy young, healthy elderly and Parkinson’ disease groups. Changes in the coupling between network metrics (in delta, theta, alpha, beta, and gamma bands) and cardiac sympathetic–vagal dynamics, and their individual mean values are presented. **Bold** indicates significance (***p*** < **0. 05**, which was confirmed by a permutation test); ***p*** < **0. 001** indicates that none of the **10**, **000** random permutations surpassed the effect magnitude from the original samples.

|  | Brain–heart coupling |  |  |  | Brain |  |
| --- | --- | --- | --- | --- | --- | --- |
| | $CSI_{HRV}$ | | $CPI_{HRV}$ | | | |
| | $E_g$ | $Q$ | $E_g$ | $Q$ | $E_g$ | $Q$ |
| $\alpha$ | $p = 0.6972$<br>$\chi^2 = 0.7561$ | $p = 0.9694$<br>$\chi^2 = 0.0601$ | $p = 0.1012$<br>$\chi^2 = 4.5244$ | $p = 0.1619$<br>$\chi^2 = 3.6289$ | **$p < 0.0001$**<br>**$\chi^2 = 38.4451$** | **$p < 0.0001$**<br>**$\chi^2 = 41.1285$** |
| $\beta$ | **$p = 0.0032$**<br>**$\chi^2 = 11.0034$** | $p = 0.6640$<br>$\chi^2 = 0.8008$ | $p = 0.3012$<br>$\chi^2 = 2.4319$ | $p = 0.1225$<br>$\chi^2 = 4.2489$ | **$p < 0.0001$**<br>**$\chi^2 = 41.4100$** | **$p < 0.0001$**<br>**$\chi^2 = 32.7429$** |
| $\gamma$ | **$p = 0.0003$**<br>**$\chi^2 = 13.9019$** | $p = 0.7340$<br>$\chi^2 = 0.6472$ | $p = 0.8708$<br>$\chi^2 = 0.2924$ | $p = 0.6463$<br>$\chi^2 = 0.9038$ | **$p < 0.0001$**<br>**$\chi^2 = 37.5683$** | **$p < 0.0001$**<br>**$\chi^2 = 22.5341$** |
| Heart |  |  |  |  |  |  |
| Sympathetic activity |  | Parasympathetic activity |  | R-R baseline |  |  |
| $CSI_{HRV}$ | $CSI$ | $CPI_{HRV}$ | $CPI$ | $CCD$ | | |
| **$p = 0.0260$**<br>**$\chi^2 = 7.2649$** | **$p = 0.0148$**<br>**$\chi^2 = 8.2867$** | $p = 0.1729$<br>$\chi^2 = 3.4243$ | $p = 0.2135$<br>$\chi^2 = 3.1281$ | **$p = 0.0014$**<br>**$\chi^2 = 12.3633$** | | |

Regarding the network analysis, metrics were found to be statistically significant across all frequency bands. Specifically, a consistent and significant trend was observed in the healthy young vs. healthy elderly and healthy young vs. PD comparisons, characterized by a progressive decrease in modularity alongside a significant increase in global efficiency across the entire frequency spectrum studied. These findings were further validated by unpaired Wilcoxon rank-sum tests, which confirmed the significant differences between groups initially identified through the Kruskal-Wallis analysis. A graphical representation is showed in Figure 2 (see Supplementary Material, Table S1 for detailed information on Wilcoxon tests results).

**Figure 2.**
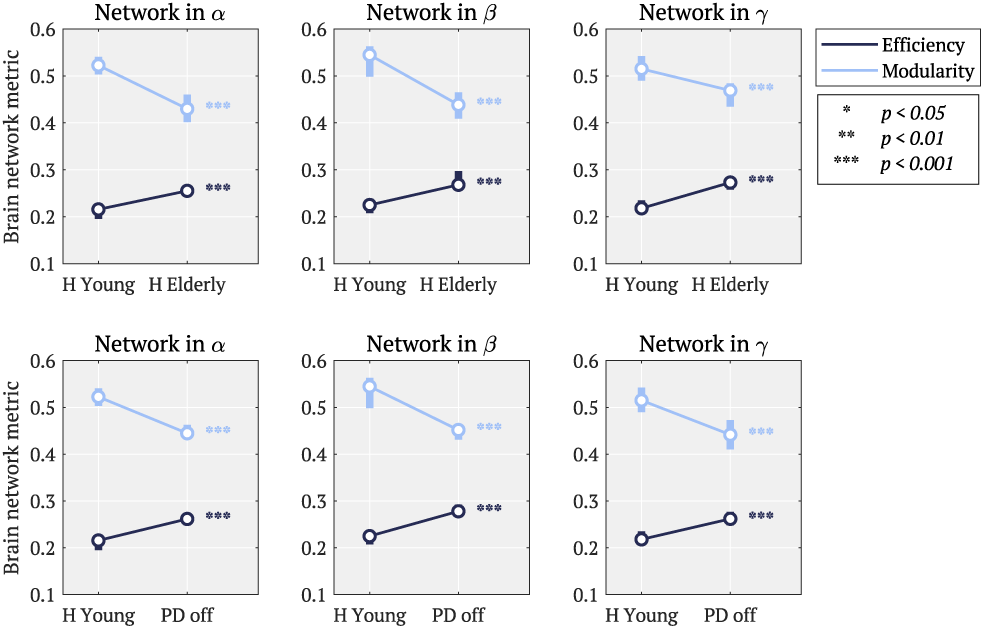
Significant differences in mean Efficiency and Modularity were observed in healthy young vs. healthy elderly and healthy young vs. PD comparisons. Data points represent the median for each group, with vertical error bars indicating the interquartile range. Dark blue bars/markers represent Efficiency, while light blue denotes Modularity. Statistical significance is indicated by asterisks (∗ ***p*** < **0. 05**, ∗∗ ***p*** < **0. 01**, ∗∗∗ ***p*** < **0. 001**); ***p*** < **0. 001** indicates that none of the **10**, **000** random permutations surpassed the effect magnitude from the original samples.

The healthy elderly group and PD groups were investigated with Spearman test to further explore the relationships between metrics and ageing. The statistical results of healthy elderly reported in Table 2 and the results of PD patients in Table 3. Ageing in healthy elderly group was found to have a significant positive correlation with increasing coupling in alpha efficiency-sympathetic activity. These analyses were not significant when testing alpha efficiency and sympathetic activity separately. However, we found a significant correlation with gamma modularity. We did not find any of these correlations with ageing in the PD group. These associations are depicted in Figure 3.

**Figure 3.**
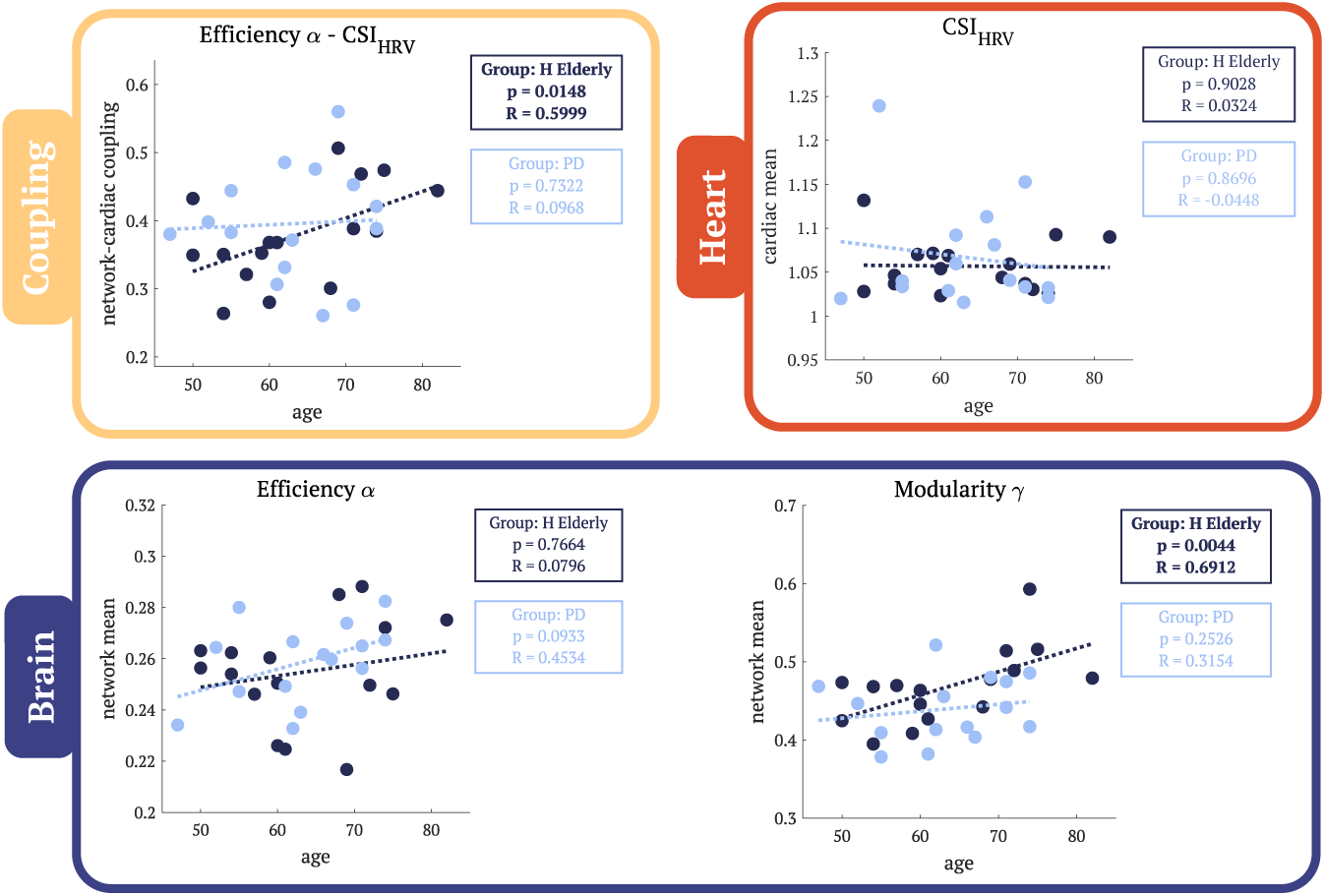
Comparison of correlation strengths across brain, heart, and coupling metrics with age. Correlations were evaluated using Spearman’s rank correlation test. In all panels, dark blue denotes the healthy elderly group, while light blue represents the Parkinson’s Disease group. Statistical results are displayed adjacent to each panel, color-coded by group. Yellow Panel (Coupling): Brain–heart Interplay indices, specifically Efficiency α and CSI_HRV_. Blue Panel (Brain): Cortical metrics independent of coupling; Efficiency α was non-significant for both groups, while Modularity γ reached significance only for the healthy elderly group. Red Panel (Heart): Isolated cardiac sympathetic variability index. **Bold** indicates significance (***p*** < **0. 05**, which was confirmed by a permutation test).

**Table 2.** Spearman correlation results on differences between brain network organization, cardiac autonomic dynamics, brain–heart interaction and ageing in Healthy Elderly group. **Bold** indicates significance (***p*** < **0. 05**, which was confirmed by a permutation test).

|  | Brain–heart coupling |  |  |  | Brain |  |
| --- | --- | --- | --- | --- | --- | --- |
| | $CSI_{HRV}$ | | $CPI_{HRV}$ | | | |
| | $E_g$ | $Q$ | $E_g$ | $Q$ | $E_g$ | $Q$ |
| $\alpha$ | **$p = 0.0148$**<br>**$R = 0.5999$** | $p = 0.2072$<br>$R = 0.3360$ | $p = 0.5878$<br>$R = -0.1444$ | $p = 0.2584$<br>$R = -0.2992$ | $p = 0.7664$<br>$R = 0.0796$ | $p = 0.9385$<br>$R = 0.0206$ |
| $\beta$ | $p = 0.2945$<br>$R = 0.2786$ | $p = 0.2085$<br>$R = 0.3301$ | $p = 0.9792$<br>$R = 0.0074$ | $p = 0.8863$<br>$R = 0.0368$ | $p = 0.9948$<br>$R = -0.0015$ | $p = 0.6628$<br>$R = 0.1120$ |
| $\gamma$ | $p = 0.7183$<br>$R = 0.0958$ | $p = 0.2339$<br>$R = 0.3139$ | $p = 0.7134$<br>$R = 0.0973$ | $p = 0.6744$<br>$R = 0.1120$ | $p = 0.8214$<br>$R = 0.0604$ | **$p = 0.0044$**<br>**$R = 0.6912$** |
| Heart |  |  |  |  |  |  |
| Sympathetic activity |  |  | Parasympathetic activity |  | R-R baseline |  |

| $CSI_{HRV}$ | $CSI$ | $CPI_{HRV}$ | $CPI$ | $CCD$ |
| --- | --- | --- | --- | --- |
| $p = 0.9024$<br>$R = 0.0324$ | $p = 0.5921$<br>$R = 0.1430$ | $p = 0.4024$<br>$R = 0.2196$ | $p = 0.3915$<br>$R = 0.2270$ | $p = 0.9386$<br>$R = 0.0206$ |

**Table 3.**
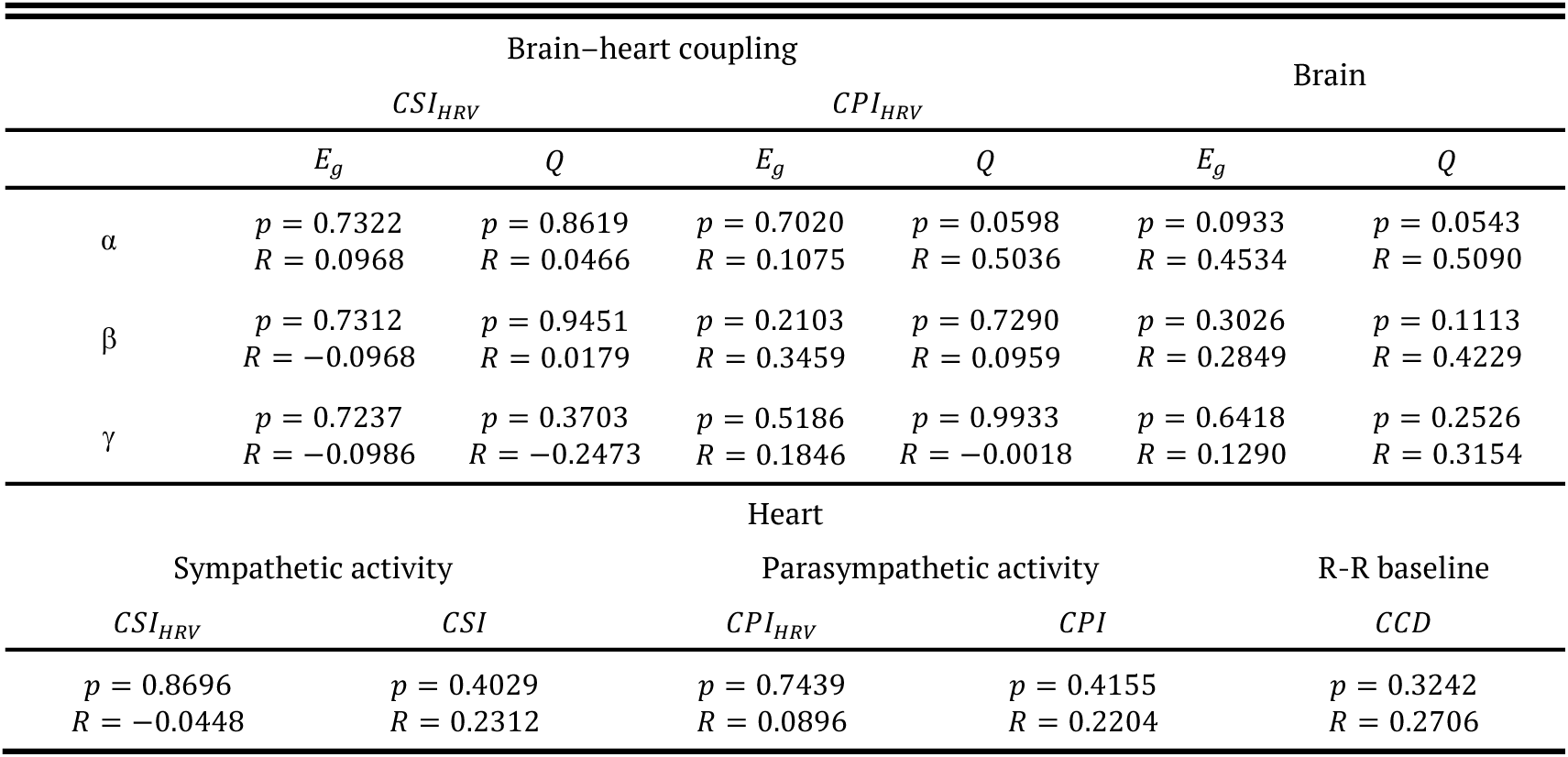
Spearman correlation results on differences between brain network organization, cardiac autonomic dynamics, brain–heart interaction and ageing in Parkinson off group. **Bold** indicates significance (***p*** < **0. 05**, which was confirmed by a permutation test).

### Cognitive ability

Both the healthy elderly and PD groups were assessed using the MMSE. MMSE scores for those groups ranged between 26 and 30, indicating no cognitive impairment across participants. We examined whether this subclinical variability could be explained with brain–heart network metrics. We did not find any correlation between the MMSE score and the brain–heart coupling, brain and heart metrics (see Table 4). Instead, we found significant correlations between the MMSE scores in the PD group, as shown in Table 5. A positive correlation appeared with the coupling between alpha modularity and cardiac dynamics, being stronger in the coupling with parasympathetic activity. In addition, a negative correlation appeared with alpha efficiency when compared separately (Figure 4). A visual representation in time of the stronger relation between coupling and MMSE is provided in Figure 5. On the right, the temporal trajectories of parasympathetic activity and alpha-band modularity are compared between two subjects with opposing MMSE scores. These findings suggest that brain–heart coupling may serve as an adequate biomarker for exploring novel aspects of the disease.

**Figure 4.**
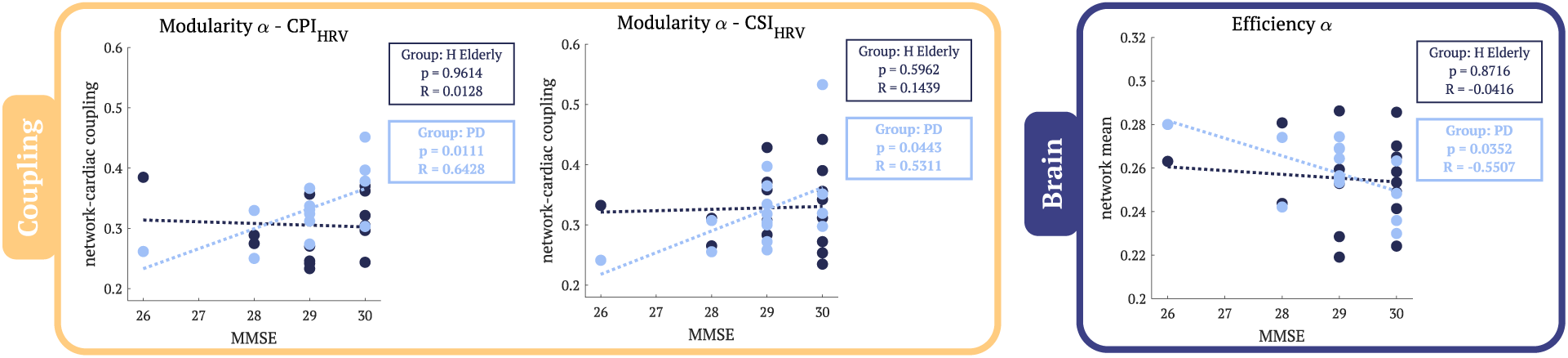
Comparison of correlation strengths across brain, heart, and coupling metrics with MMSE. Correlations were evaluated using Spearman’s rank correlation test. In all panels, dark blue denotes the Healthy Elderly (HE) group, while light blue represents the Parkinson’s Disease (PD) group. Statistical results are displayed adjacent to each panel, color-coded by group. Yellow Panel (Coupling): Brain–heart Interaction indices, specifically Modularity α-CPI_HRV_ and Modularity α-CSI_HRV_. Blue Panel (Brain): Modularity α was non-significant for both groups, while Efficiency α and Modularity δ reached significance only for the PD group. **Bold** indicates significance (***p*** < **0. 05**, which was confirmed by a permutation test).

**Figure 5.**
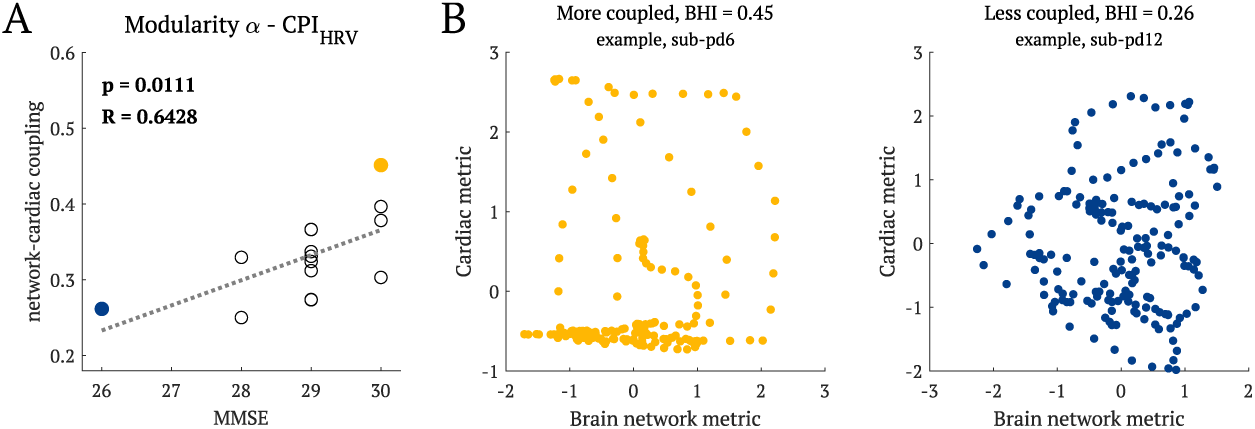
Panel A: The scatter plot illustrates the strongest correlation identified between the PD group metrics and MMSE scores, consistent with the results presented in Figure 4. Panel B: Phase-space representation of normalized temporal patterns of neural network on x-axis and autonomic metrics on y-axis for two representative cases. Subject 6 (yellow) demonstrates the strongest coupling, corresponding to a higher MMSE score, while Subject 12 (blue) exhibits lower coupling, associated with a lower MMSE score.

**Table 4.**
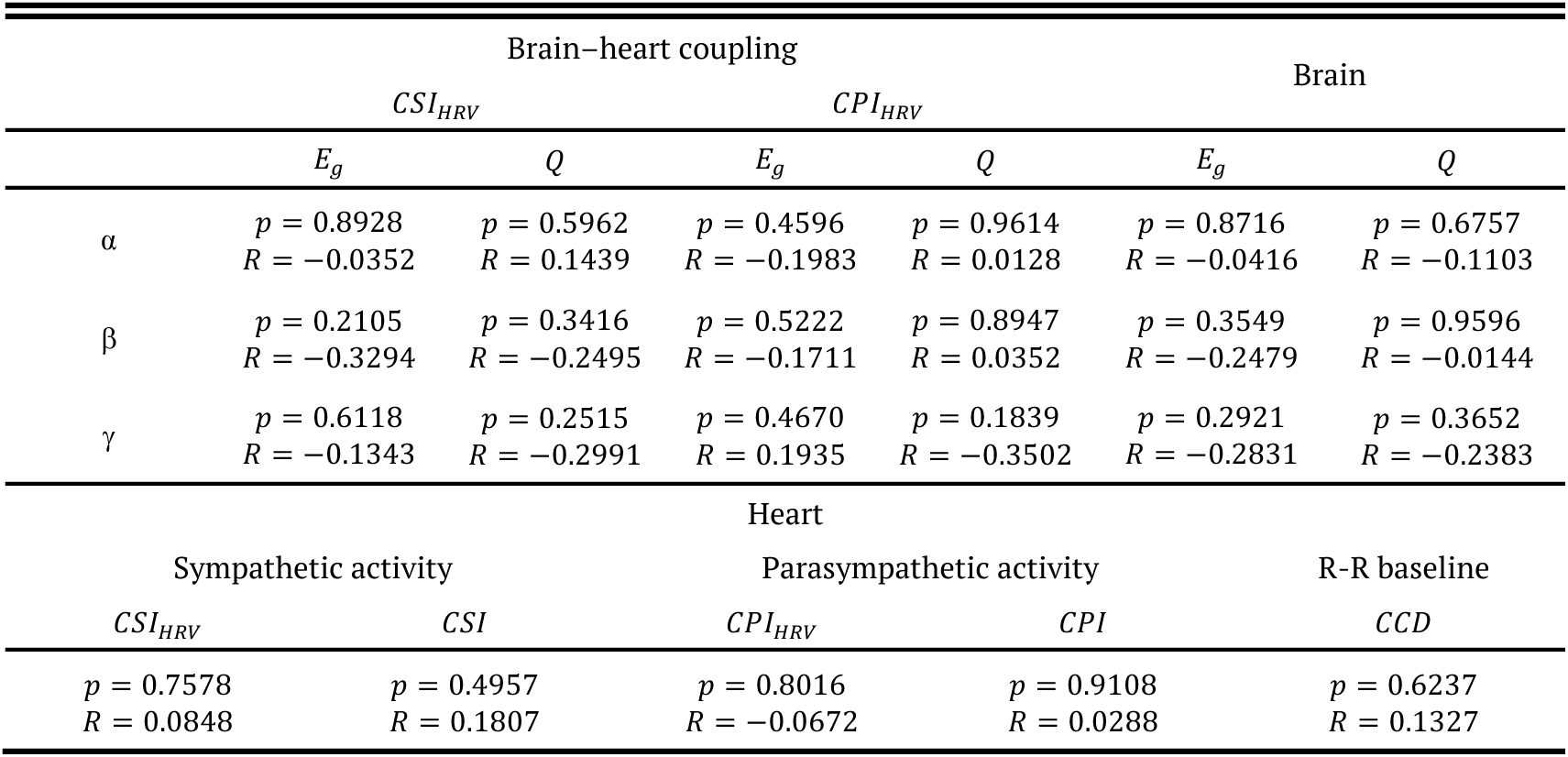
Spearman correlation results on differences between brain network organization, cardiac autonomic dynamics, brain–heart interaction and cognitive ability in Healthy Elderly group. **Bold** indicates significance (***p*** < **0. 05**, which was confirmed by a permutation test).

**Table 5.**
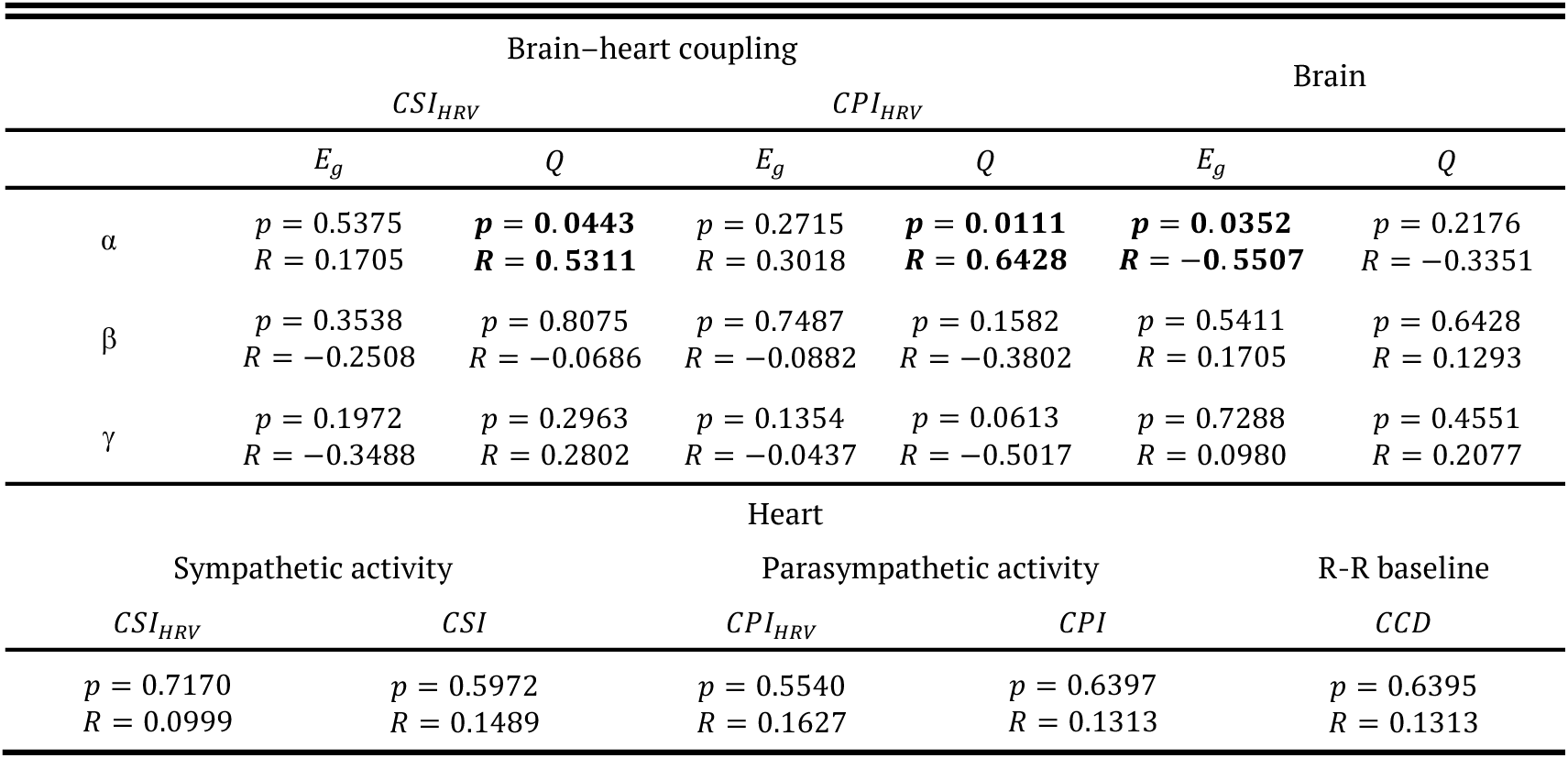
Spearman correlation results on differences between brain network organization, cardiac autonomic dynamics, brain–heart interaction and cognitive ability in Parkinson disease group. **Bold** indicates significance (***p*** < **0. 05**, which was confirmed by a permutation test).

**Table 6.** Summary of trial distribution for the FOG dataset following the subject selection pipeline. ‘Task” denotes the number of walking tests completed by each subject, consistent with the terminology used in the original open-source dataset.

| Trial | Subject | Task | Pre-FOG duration | FOG duration |
| --- | --- | --- | --- | --- |
| 1 | 10 | 2 | 58 s | 44 s |
| 2 | 10 | 2 | 50 s | 56 s |
| 3 | 12 | 4 | 60 s | 74 s |
| 4 | 12 | 4 | 68 s | 24 s |
| 5 | 8 | 1 | 40 s | 24 s |
| 6 | 8 | 2 | 48 s | 71 s |

### Freezing of gait

We identified a total of 6 trials accomplishing with the selection criteria of FOG events. Specifically, trials 1 and 2 were recorded from subject 10, trials 3 and 4 from subject 12, and trials 5 and 6 from subject 8. Further details are reported in Table 7.

For each trial, brain network, autonomic dynamics and brain–heart interaction metrics were calculated. The analysis specifically focused on Global Efficiency and Modularity within the beta band and the overall effect of HR and HRV in the two autonomic branches. This choice was motivated by the fact that subjects were engaged in a walking task, and the beta rhythm is well-documented to be closely associated with motor control and movement dynamics. Moreover, it is preferred to inspect the sum effect of the sympathetic and parasympathetic branch because subjects were not in resting state and it is important to take into account the variations in their general effect.

To investigate potential trends in the coupling during the transition from the period preceding the FOG onset to the actual FOG episode, the Maximal Information Coefficient and the mean values of both brain and heart metrics were computed over two distinct intervals: the pre-FOG window (from −10s to 0s) and the FOG window (from 0s to +10s). Figure 6 illustrates the transitions in brain–heart coupling estimation and mean individual metrics between the pre-FOG and FOG windows. For all metrics, the transition between the two states was quantified by calculating the difference between the pre-FOG and FOG onset values. According to this convention, negative values denote an increase in the parameter’s magnitude at the onset of FOG. Panels A and B display the variation in the MIC (Δ*MIC*) calculated between beta Modularity and parasympathetic activity, over the corresponding changes in isolated brain and heart components. Similarly, Panels C and D show the relationships concerning the coupling between beta Efficiency and sympathetic activity. Our results revealed a distinct cluster for the coupling indices, which appear to either increase or remain stable during the transition regardless of whether the individual brain or heart metrics increase or decrease. Finally, Panel E provides representative temporal traces from two subjects, highlighting the contrast between stable MIC (yellow) and increased MIC (green) during the FOG event.

**Figure 6.**
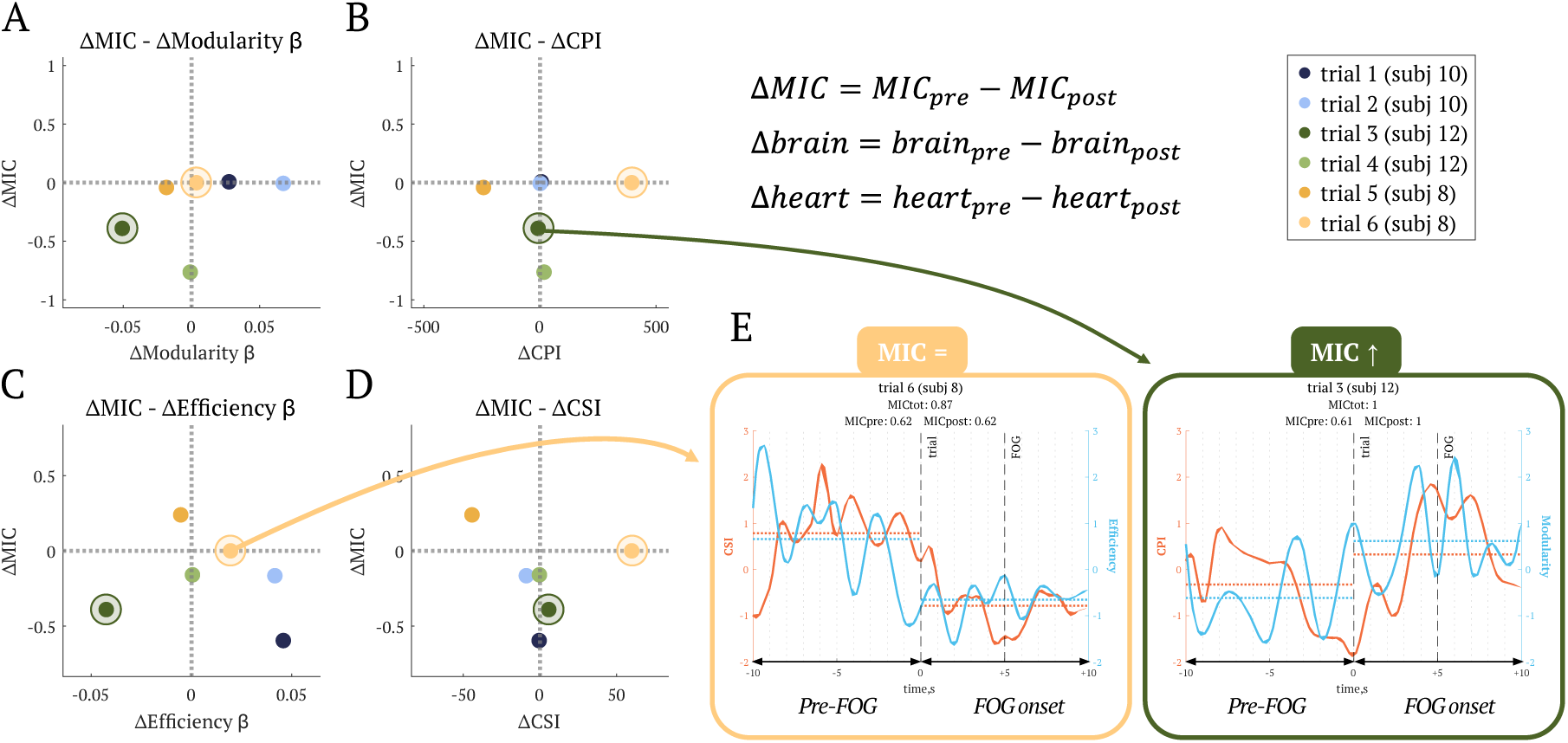
Graphical representation of brain–heart coupling transitions with FOG onset. Panels A–D: Metric changes calculated as Δ***metric*** = ***metric***_***p******re*** - ***FOG***_ − ***metric***_***FOG***_, where ***metric*** can be either coupling, mean value of brain or mean value of heart in the pre-FOG and FOG window. Panels A-B show MIC variation between β Modularity and global parasympathetic activity over variation in β Modularity (A) or CPI (B); Panels C-D show βEfficiency and global sympathetic coupling over variation in βEfficiency (C) or CSI (D). (E) Representative temporal traces showing stable (yellow) and increased (green) MIC during FOG episodes.

## Discussion

Our results indicate that the study of brain–heart interactions represents a promising framework for investigating large-scale physiological alterations associated with neurodegeneration and healthy ageing. Brain–heart interaction metrics have demonstrated discriminatory power comparable to, and in some cases greater than, approaches based solely on brain network organization or cardiac autonomic dynamics.

The comparison between healthy young, healthy elderly, and PD groups highlights the capacity of brain–heart interactions to display differences across various physiological and pathological conditions. Significant differences in coupling were identified specifically for beta efficiency-sympathetic activity and gamma efficiency-sympathetic activity. Brain features showed the strongest discrimination; however, brain–heart interaction metrics exhibited superior capacity compared to stand-alone cardiac indices.

During ageing, alterations occur in the cardiovascular and nervous systems as adaptive responses to internal changes and environmental challenges. It is well established that resting HRV declines with age, primarily due to impaired sympatho-vagal modulation [54], [55]. Our findings support this age-related reduction in autonomic regulation, as reflected by significant alterations in sympathetic activity [56]. Although elevated HRV is generally interpreted as a marker of enhanced vagal modulation, it is also associated with higher physical fitness levels, suggesting that lifestyle-related factors may substantially influence HRV measures across ageing. This potential confounding effect supports the hypothesis that brain network organization and brain–heart interaction metrics may provide more robust biomarkers of physiological ageing than isolated HRV indices alone.

In the context of brain network interaction, it has been reported that a higher HRV was linked to stronger network centrality in several brain regions [54]. Moreover, accumulating evidence indicates that alterations in intrinsic brain activity represent a key feature of normal brain ageing, leading to age-related functional reorganization and consequent changes in resting-state network architecture. This framework may also explain why the effects observed in the present study appear more pronounced at the neural level, suggesting that the detected differences in brain–heart coupling are primarily driven by changes in brain dynamics. Across measures of brain network organization, significant differences were observed between healthy young participants and both healthy elderly and PD groups. In all frequency bands, healthy elderly and PD participants exhibited patterns consistent with increased functional integration and reduced segregation. These findings support the hypothesis that ageing is associated with neural de-differentiation [57], [58]. More specifically, the brain may progressively shift from highly specialized small-world networks toward a more diffuse and less efficient topology [46], [59]. This transition is thought to reflect the global decline in gray matter density associated with ageing. Structural deterioration may therefore promote compensatory recruitment of broader functional networks, resulting in more diffuse connectivity patterns. However, such compensatory recruitment is not necessarily associated with preserved performance [60] and may instead reflect reduced neural efficiency.

We investigated the potential relationship between the degree of cognitive ability, as measured by MMSE, and the various metrics examined in this study. A comparison of these correlations between healthy elderly and PD groups highlights a distinct divergence between the two populations. A positive correlation was observed between higher cognitive performance and enhanced coupling between alpha-band modularity and both sympathetic and parasympathetic indices. While we did not observe associations between brain–heart metrics and chronological age in PD patients, we found that increased global hyper-synchronicity within the alpha band was significantly correlated with lower MMSE scores. These findings suggest that brain–heart interactions may capture individual differences in cognitive vulnerability in PD rather than chronological ageing itself, potentially reflecting early decline in general cognitive functioning. Nevertheless, it is important to remark that elderly participants in this study did not fall within the clinical MMSE range for a Mild Cognitive Impairment diagnosis [39]. Consequently, it remains uncertain whether the observed correlation strength would be maintained in patients with overt cognitive decline.

MMSE scores and network metrics were found to have significant correlations exclusively within the PD group, but the relationship was inversely proportional in this instance. These results align with previous findings that characterize increased global synchronization as a compensatory, yet often maladaptive, response to the loss of local complexity in Mild Cognitive Impairment patients [46], [61], [62]. Specifically, it has been shown that progressive Mild Cognitive Impairment patients exhibit higher alpha-band synchronization between the right anterior cingulate and temporo-occipital regions compared to stable subjects. This hyper synchronization was found to be inversely correlated with cognitive performance, as well as with the volumes of brain regions [61]. The increase in global synchronization and long-range connectivity is often interpreted as a compensatory response to the loss of local complexity. This change results in a higher metabolic energy expenditure and a transition toward a more random, less efficient network structure compared to healthy controls [46]. These findings likely emerged exclusively within the PD group because the pathology itself is fundamentally characterized by an abnormal increase in neural synchrony [4]. Consequently, these network alterations and their impact on cognitive ability are more pronounced and detectable in PD patients than in healthy subjects.

Although it is well-established that resting-state brain oscillations shift with ageing and influence cognitive performance [63], scientific literature remains inconsistent regarding specific changes in EEG power related to cognitive ability. As discussed by Cesnaite et al. [63], these discrepancies are not random but stem from specific methodological aspects: from considering rhythmic and non-rhythmic components, to variability in frequency band definitions, which can lead to inconsistent measurements or make age-related differences disappear. Brain–heart interaction frameworks offer an alternative to those measurements by focusing on the synergy between physiological systems, rather than specific increases or decreases of a defined marker.

With respect to our analyses in FOG, in the transition from the pre-FOG to the FOG onset condition, a cluster was identified in the change of brain–heart interactions. Within this shift, the coupling magnitude appeared to either remain stable or increase, regardless of the mean trends observed in the individual heart and brain metrics from which the coupling was derived. Among the six identified trials, most exhibited an increase in coupling strength upon entering the FOG state. FOG onset likely emerges from a complex systemic failure involving motor, emotional, and cognitive processes [5], [6], [10], [11], [12], [13], [32], [47], [48], [49]. One possible interpretation is that excessive crosstalk among physiological networks, including motor and limbic circuits, overloads the dopamine-depleted striatum and disrupts brainstem locomotor control [6], [48]. This could manifest as a failure in the beta suppression, typically observed in movement initiation [6], [47]. In line with this, synchronized beta bursts can be found during FOG episodes [6], [48], which can occur across multiple regions, spanning both subcortical structures, such as the subthalamic nucleus and the basal ganglia [6], [47], and cortical areas, including the supplementary motor area, primary motor cortex, and somatosensory cortex [5], [47]. Additionally, an increase in local connectivity has been observed immediately prior to FOG onset within the visual areas and the right frontoparietal cortex, among others [49]. These findings suggest that FOG-associated brain dynamics involve local and distributed changes.

Patients prone to FOG exhibit low resting HRV, signaling a potential imbalance between the sympathetic and parasympathetic branches, sometimes interpreted as a simultaneous co-inhibition of both autonomic branches [12], [13]. Although the changes in HRV during FOG episodes can vary, they are typically associated with increases in heart rate [11], [32]. These dynamics could be triggered by increases in sympathetic activity likely driven by the limbic system in response to emotional stress. This heart rate increase is also attributed to the physiological response to effort and intention to move, despite motor failure [10], [11], and to the compensatory hyperactivity of the pedunculopontine nucleus during FOG [12]. Furthermore, proper recruitment of the cuneiform nucleus is associated with higher HRV and a lower incidence of freezing [13]. Our results regarding the variation of cardiac sympathetic and parasympathetic indices appear to be highly subject-dependent. While clear changes are observed during the transition between conditions, they do not exhibit a consistent trend comparable to the brain network metrics, and the overall differences remain relatively small. The presence of such anomalies warrants further investigation to better isolate the impact of individual variability on cardiovascular responses during FOG. Our findings suggest that coupling tends to increase or remain stable between the specific brain and heart components during FOG episodes. Specifically, this strengthening of coupling is observed between global network integration and the sympathetic branch, as well as between network segregation and the parasympathetic branch. Given that FOG can be triggered by a variety of factors, it is probable that the episodes analyzed in this study originated from different causes, each potentially influencing the heart-brain interaction in unique ways. It is important to note that, among all the FOG episodes studied, the increases in brain–heart coupling were not associated with consistent increases or decreases in brain and heart standalone markers. Future research would benefit to conduct post-episode patient assessments to evaluate the effect of their subjective experiences, such as fear, anxiety, or cognitive stress, with specific trends in the coupling metrics. Furthermore, investigating the opposite relationship could provide equally insightful information by examining how FOG termination mechanisms impact neural network reorganization and autonomic responses.

Future studies would benefit from larger cohorts to better capture the complex trajectories in ageing and PD, but also with the inclusion of other measurements, such as hemodynamics [64]. Inclusions could a wider range of symptoms manifestations, including those with diagnosed cognitive impairment. Additional inclusion criteria could account for the specific socio-demographic context and other risk factors of the participants [65]. These adjustments are particularly necessary in highly educated cohorts, where the tests may fail to detect early cognitive decline due to high baseline performance [40], [66]. In the FOG dataset, a significant number of recordings had to be excluded during the subject selection phase in the dataset utilized for the FOG analysis, where strict inclusion criteria were necessary to ensure signal integrity. Indeed, only six FOG events across twelve subjects were suitable for the final analysis. This limited sample size is attributable to the high frequency of closely spaced, short-duration FOG episodes. A rigorous selection was essential, as the proximity of consecutive events made it impossible to guarantee that the pathophysiological FOG mechanisms were triggered from a standard gait pattern. The exclusion of such episodes ensured a more accurate calculation of brain–heart interaction metrics by avoiding the impact of a previous event that could act as a confounding factor. Furthermore, several subjects had to be excluded due to poor ECG signal quality. In their study, Zhang et al. [31] successfully induced FOG; however, the experimental protocols appeared to be in some cases overly complex, leading to continuous freezing episodes. Despite the limited sample size, this work represents one of first endeavors multiple aspects of PD within a framework of brain–heart interactions. Autonomic deficits lead to multifaceted clinical issues, including reduced HRV and diminished synchronization between cardiac activity and specific cortical regions [3], [67]. Changes in the interplay between brain and heart is not exclusive of conditions involving dysautonomia [17], [21], [68], [69], [70], [71]..

Our results demonstrate the feasibility of the proposed pipeline, establishing a robust foundation for its application in future, larger-scale and dedicated clinical studies. The development and refinement of these new brain-body frameworks are essential to provide a more comprehensive understanding of the insights into the role of these interactions in both health and disease. Such a holistic perspective opens new possibilities for development of more precise, multi-modal biomarkers for PD and manifestation associated with ageing.

## Conclusion

By comparing various metrics, this study demonstrated that manifestations of ageing and PD can be captured by measures of brain–heart interactions, including clinical and subclinical conditions. These findings add to the growing literature confirming the importance of monitoring brain-body interactions in neurological disorders. Our analyses revealed previously unexplored aspects of both pathological and physiological states. Such a framework may help clarify whether cognitive impairment and normal ageing share common physiological mechanisms or instead represent distinct processes. Moreover, understanding the dynamics of this biomarker could provide crucial insights into the mechanisms driving the escalation of FOG in PD. Ultimately, the study of brain–heart interactions represents a powerful tool for addressing large-scale physiological networks in highly heterogeneous conditions such as PD, opening the way for more personalized monitoring strategies. Our findings offer promising new perspectives for the application of this analytical pipeline in future research.

## Data Availability

All data produced in the present study are available upon reasonable request to the authors

## References

[1] S. Sveinbjornsdottir, “The clinical symptoms of Parkinson’s disease,” J. Neurochem., vol. 139, no. S1, pp. 318–324, Oct. 2016, doi: 10.1111/jnc.13691.

[2] J. S. George, J. Strunk, R. Mak-McCully, M. Houser, H. Poizner, and A. R. Aron, “Dopaminergic therapy in Parkinson’s disease decreases cortical beta band coherence in the resting state and increases cortical beta band power during executive control,” NeuroImage Clin., vol. 3, pp. 261–270, 2013, doi: 10.1016/j.nicl.2013.07.013.

[3] A. Criscuolo, A. Pelucchi, M. Schwartze, and S. A. Kotz, “A systematic review on autonomic dysfunctions in Parkinson’s disease,” May 12, 2025, In Review. doi: 10.21203/rs.3.rs-6621647/v1.

[4] C. Hammond, H. Bergman, and P. Brown, “Pathological synchronization in Parkinson’s disease: networks, models and treatments,” Trends Neurosci., vol. 30, no. 7, pp. 357–364, Jul. 2007, doi: 10.1016/j.tins.2007.05.004.

[5] N. Madetko-Alster et al., “The role of the somatosensory cortex in self-paced movement impairment in Parkinson’s disease,” Clin. Neurophysiol., vol. 171, pp. 11–17, Mar. 2025, doi: 10.1016/j.clinph.2025.01.001.

[6] J. S. Marquez et al., “Neural Correlates of Freezing of Gait in Parkinson’s Disease: An Electrophysiology Mini-Review,” Front. Neurol., vol. 11, p. 571086, Nov. 2020, doi: 10.3389/fneur.2020.571086.

[7] S. Jain, “Multi-organ autonomic dysfunction in Parkinson disease,” Parkinsonism Relat. Disord., vol. 17, no. 2, pp. 77–83, Feb. 2011, doi: 10.1016/j.parkreldis.2010.08.022.

[8] K. Wakabayashi and H. Takahashi, “Neuropathology of Autonomic Nervous System in Parkinson’s Disease,” Eur. Neurol., vol. 38, no. 2, pp. 2–7, 1997, doi: 10.1159/000113469.

[9] M. Mather, “Autonomic dysfunction in neurodegenerative disease,” Nat. Rev. Neurosci., vol. 26, no. 5, pp. 276–292, May 2025, doi: 10.1038/s41583-025-00911-8.

[10] H. Cockx, J. Nonnekes, B. R. Bloem, R. Van Wezel, I. Cameron, and Y. Wang, “Dealing with the heterogeneous presentations of freezing of gait: how reliable are the freezing index and heart rate for freezing detection?,” J. NeuroEngineering Rehabil., vol. 20, no. 1, p. 53, Apr. 2023, doi: 10.1186/s12984-023-01175-y.

[11] I. Maidan, M. Plotnik, A. Mirelman, A. Weiss, N. Giladi, and J. M. Hausdorff, “Heart rate changes during freezing of gait in patients with Parkinson’s disease,” Mov. Disord., vol. 25, no. 14, pp. 2346–2354, Oct. 2010, doi: 10.1002/mds.23280.

[12] B. Heimler et al., “Heart-rate variability as a new marker for freezing predisposition in Parkinson’s disease,” Parkinsonism Relat. Disord., vol. 113, p. 105476, Aug. 2023, doi: 10.1016/j.parkreldis.2023.105476.

[13] J. Y. Liao and N. I. Bohnen, “Is heart rate variability the heart of the matter for freezing of gait?,” Parkinsonism Relat. Disord., vol. 113, p. 105763, Aug. 2023, doi: 10.1016/j.parkreldis.2023.105763.

[14] Y. Sharabi, G. D. Vatine, and A. Ashkenazi, “Parkinson’s disease outside the brain: targeting the autonomic nervous system,” Lancet Neurol., vol. 20, no. 10, pp. 868–876, Oct. 2021, doi: 10.1016/S1474-4422(21)00219-2.

[15] T. H. Haapaniemi, “Ambulatory ECG and analysis of heart rate variability in Parkinson’s disease,” J. Neurol. Neurosurg. Psychiatry, vol. 70, no. 3, pp. 305–310, Mar. 2001, doi: 10.1136/jnnp.70.3.305.

[16] M. A. Samuels, “The Brain–Heart Connection,” Circulation, vol. 116, no. 1, pp. 77–84, Jul. 2007, doi: 10.1161/CIRCULATIONAHA.106.678995.

[17] S. Fang and W. Zhang, “Heart–Brain Axis: A Narrative Review of the Interaction between Depression and Arrhythmia,” Biomedicines, vol. 12, no. 8, p. 1719, Aug. 2024, doi: 10.3390/biomedicines12081719.

[18] A. Silvani, G. Calandra-Buonaura, R. A. L. Dampney, and P. Cortelli, “Brain–heart interactions: physiology and clinical implications,” Philos. Trans. R. Soc. Math. Phys. Eng. Sci., vol. 374, no. 2067, p. 20150181, May 2016, doi: 10.1098/rsta.2015.0181.

[19] P. Borghammer and N. Van Den Berge, “Brain-First versus Gut-First Parkinson’s Disease: A Hypothesis,” J. Park. Dis., vol. 9, no. s2, Oct. 2019, doi: 10.3233/JPD-191721.

[20] J.-A. Palma and H. Kaufmann, “Autonomic disorders predicting Parkinson’s disease,” Parkinsonism Relat. Disord., vol. 20, pp. S94–S98, Jan. 2014, doi: 10.1016/S1353-8020(13)70024-5.

[21] D. Candia-Rivera, M. Vidailhet, M. Chavez, and F. De Vico Fallani, “A framework for quantifying the coupling between brain connectivity and heartbeat dynamics: Insights into the disrupted network physiology in Parkinson’s disease,” Hum. Brain Mapp., vol. 45, no. 5, p. e26668, Apr. 2024, doi: 10.1002/hbm.26668.

[22] D. Candia-Rivera, M. Chavez, and F. De Vico Fallani, “Measures of the coupling between fluctuating brain network organization and heartbeat dynamics,” Netw. Neurosci., vol. 8, no. 2, pp. 557–575, Jul. 2024, doi: 10.1162/netn_a_00369.

[23] D. Candia-Rivera and F. de V. Fallani, “Robust and time-resolved estimation of cardiac sympathetic and parasympathetic indices”.

[24] A. P. Rockhill, Nicko Jackson, Jobi George, A. Aron, and N. C. Swann, “UC San Diego Resting State EEG Data from Patients with Parkinson’s Disease.” Openneuro, 2022. doi: 10.18112/OPENNEURO.DS002778.V1.0.5.

[25] S. Appelhoff et al., “MNE-BIDS: Organizing electrophysiological data into the BIDS format and facilitating their analysis,” J. Open Source Softw., vol. 4, no. 44, p. 1896, Dec. 2019, doi: 10.21105/joss.01896.

[26] N. Jackson, S. R. Cole, B. Voytek, and N. C. Swann, “Characteristics of Waveform Shape in Parkinson’s Disease Detected with Scalp Electroencephalography,” eneuro, vol. 6, no. 3, p. ENEURO.0151-19.2019, May 2019, doi: 10.1523/ENEURO.0151-19.2019.

[27] C. R. Pernet et al., “EEG-BIDS, an extension to the brain imaging data structure for electroencephalography,” Sci. Data, vol. 6, no. 1, p. 103, Jun. 2019, doi: 10.1038/s41597-019-0104-8.

[28] N. C. Swann, C. De Hemptinne, A. R. Aron, J. L. Ostrem, R. T. Knight, and P. A. Starr, “Elevated synchrony in P arkinson disease detected with electroencephalography,” Ann. Neurol., vol. 78, no. 5, pp. 742–750, Nov. 2015, doi: 10.1002/ana.24507.

[29] S. Koelstra et al., “DEAP: A Database for Emotion Analysis ;Using Physiological Signals,” IEEE Trans. Affect. Comput., vol. 3, no. 1, pp. 18–31, Jan. 2012, doi: 10.1109/T-AFFC.2011.15.

[30] H. Li, “Multimodal Dataset of Freezing of Gait in Parkinson’s Disease.” Mendeley, Jan. 26, 2021. doi: 10.17632/R8GMBTV7W2.3.

[31] W. Zhang et al., “Multimodal Data for the Detection of Freezing of Gait in Parkinson’s Disease,” Sci. Data, vol. 9, no. 1, p. 606, Oct. 2022, doi: 10.1038/s41597-022-01713-8.

[32] D. Candia-Rivera and M. Chavez, “Freezing of gait in Parkinson’s disease increases sympathetic and parasympathetic indices,” in 2024 13th Conference of the European Study Group on Cardiovascular Oscillations (ESGCO), Oct. 2024, pp. 1–2. doi: 10.1109/ESGCO63003.2024.10767055.

[33] R. Oostenveld, P. Fries, E. Maris, and J.-M. Schoffelen, “FieldTrip: Open Source Software for Advanced Analysis of MEG, EEG, and Invasive Electrophysiological Data,” Comput. Intell. Neurosci., vol. 2011, pp. 1–9, 2011, doi: 10.1155/2011/156869.

[34] D. Candia-Rivera, V. Catrambone, and G. Valenza, “The role of electroencephalography electrical reference in the assessment of functional brain–heart interplay: From methodology to user guidelines,” J. Neurosci. Methods, vol. 360, p. 109269, Aug. 2021, doi: 10.1016/j.jneumeth.2021.109269.

[35] B. Molavi and G. A. Dumont, “Wavelet-based motion artifact removal for functional near-infrared spectroscopy,” Physiol. Meas., vol. 33, no. 2, pp. 259–270, Jan. 2012, doi: 10.1088/0967-3334/33/2/259.

[36] N. P. Castellanos and V. A. Makarov, “Recovering EEG brain signals: artifact suppression with wavelet enhanced independent component analysis,” J. Neurosci. Methods, vol. 158, no. 2, pp. 300–312, Dec. 2006, doi: 10.1016/j.jneumeth.2006.05.033.

[37] J. Moeyersons, M. Amoni, S. Van Huffel, R. Willems, and C. Varon, “R-DECO: an open-source Matlab based graphical user interface for the detection and correction of R-peaks,” PeerJ Comput. Sci., vol. 5, p. e226, Oct. 2019, doi: 10.7717/peerj-cs.226.

[38] J. Pan and W. J. Tompkins, “A Real-Time QRS Detection Algorithm,” IEEE Trans. Biomed. Eng., vol. BME-32, no. 3, pp. 230–236, Mar. 1985, doi: 10.1109/TBME.1985.325532.

[39] T. N. Tombaugh and N. J. McIntyre, “The Mini-Mental State Examination: A Comprehensive Review,” J. Am. Geriatr. Soc., vol. 40, no. 9, pp. 922–935, Sep. 1992, doi: 10.1111/j.1532-5415.1992.tb01992.x.

[40] S. E. O’Bryant et al., “Detecting Dementia With the Mini-Mental State Examination in Highly Educated Individuals,” Arch. Neurol., vol. 65, no. 7, Jul. 2008, doi: 10.1001/archneur.65.7.963.

[41] F. De Vico Fallani, V. Latora, and M. Chavez, “A Topological Criterion for Filtering Information in Complex Brain Networks,” PLOS Comput. Biol., vol. 13, no. 1, p. e1005305, Jan. 2017, doi: 10.1371/journal.pcbi.1005305.

[42] G. C. Carter, “Coherence and time delay estimation,” Proc. IEEE, vol. 75, no. 2, pp. 236–255, 1987, doi: 10.1109/PROC.1987.13723.

[43] Z. Dai and Y. He, “Disrupted structural and functional brain connectomes in mild cognitive impairment and Alzheimer’s disease,” Neurosci. Bull., vol. 30, no. 2, pp. 217–232, Apr. 2014, doi: 10.1007/s12264-013-1421-0.

[44] M. Rubinov and O. Sporns, “Complex network measures of brain connectivity: Uses and interpretations,” Comput. Models Brain, vol. 52, no. 3, pp. 1059–1069, Sep. 2010, doi: 10.1016/j.neuroimage.2009.10.003.

[45] M. Conti et al., “Brain Functional Connectivity in de novo Parkinson’s Disease Patients Based on Clinical EEG,” Front. Neurol., vol. 13, p. 844745, Mar. 2022, doi: 10.3389/fneur.2022.844745.

[46] J. M. Buldú et al., “Reorganization of Functional Networks in Mild Cognitive Impairment,” PLoS ONE, vol. 6, no. 5, p. e19584, May 2011, doi: 10.1371/journal.pone.0019584.

[47] F. Karimi, J. Niu, K. Gouweleeuw, Q. Almeida, and N. Jiang, “Movement-related EEG signatures associated with freezing of gait in Parkinson’s disease: an integrative analysis,” Brain Commun., vol. 3, no. 4, p. fcab277, Oct. 2021, doi: 10.1093/braincomms/fcab277.

[48] C. Anidi et al., “Neuromodulation targets pathological not physiological beta bursts during gait in Parkinson’s disease,” Neurobiol. Dis., vol. 120, pp. 107–117, Dec. 2018, doi: 10.1016/j.nbd.2018.09.004.

[49] Y. Tian, E. Matar, S. Berkovsky, and S. J. G. Lewis, “Aberrant Beta-Band Network Alteration Preceding Freezing of Gait in Parkinson’s Disease,” Mov. Disord., p. mds.70230, Feb. 2026, doi: 10.1002/mds.70230.

[50] D. Candia-Rivera, V. Catrambone, R. Barbieri, and G. Valenza, “Integral pulse frequency modulation model driven by sympathovagal dynamics: Synthetic vs. real heart rate variability,” Biomed. Signal Process. Control, vol. 68, p. 102736, Jul. 2021, doi: 10.1016/j.bspc.2021.102736.

[51] D. Candia-Rivera, L. Faes, F. D. V. Fallani, and M. Chavez, “Measures and Models of Brain–heart Interactions,” IEEE Rev. Biomed. Eng., vol. 19, pp. 24–40, 2026, doi: 10.1109/RBME.2025.3529363.

[52] D. N. Reshef et al., “Detecting Novel Associations in Large Data Sets,” Science, vol. 334, no. 6062, pp. 1518–1524, Dec. 2011, doi: 10.1126/science.1205438.

[53] D. Candia-Rivera and G. Valenza, “Cluster permutation analysis for EEG series based on non-parametric Wilcoxon–Mann–Whitney statistical tests,” SoftwareX, vol. 19, p. 101170, 2022, doi: 10.1016/j.softx.2022.101170.

[54] D. Kumral et al., “The age-dependent relationship between resting heart rate variability and functional brain connectivity,” NeuroImage, vol. 185, pp. 521–533, Jan. 2019, doi: 10.1016/j.neuroimage.2018.10.027.

[55] R. E. De Meersman and P. K. Stein, “Vagal modulation and aging,” Biol. Psychol., vol. 74, no. 2, pp. 165–173, Feb. 2007, doi: 10.1016/j.biopsycho.2006.04.008.

[56] R. Parashar, “Age Related Changes in Autonomic Functions,” J. Clin. Diagn. Res., 2016, doi: 10.7860/JCDR/2016/16889.7497.

[57] K. Cassady et al., “Sensorimotor network segregation declines with age and is linked to GABA and to sensorimotor performance,” NeuroImage, vol. 186, pp. 234–244, Feb. 2019, doi: 10.1016/j.neuroimage.2018.11.008.

[58] M. Y. Chan, D. C. Park, N. K. Savalia, S. E. Petersen, and G. S. Wig, “Decreased segregation of brain systems across the healthy adult lifespan,” Proc. Natl. Acad. Sci. U. S. A., vol. 111, no. 46, pp. E4997–5006, Nov. 2014, doi: 10.1073/pnas.1415122111.

[59] D. Meunier, E. A. Stamatakis, and L. K. Tyler, “Age-related functional reorganization, structural changes, and preserved cognition,” Neurobiol. Aging, vol. 35, no. 1, pp. 42–54, Jan. 2014, doi: 10.1016/j.neurobiolaging.2013.07.003.

[60] L. K. Tyler, P. Wright, B. Randall, W. D. Marslen-Wilson, and E. A. Stamatakis, “Reorganization of syntactic processing following left-hemisphere brain damage: does right-hemisphere activity preserve function?,” Brain, vol. 133, no. 11, pp. 3396–3408, Nov. 2010, doi: 10.1093/brain/awq262.

[61] M. E. López et al., “Alpha-Band Hypersynchronization in Progressive Mild Cognitive Impairment: A Magnetoencephalography Study,” J. Neurosci., vol. 34, no. 44, pp. 14551–14559, Oct. 2014, doi: 10.1523/JNEUROSCI.0964-14.2014.

[62] H.-C. Baggio et al., “Cognitive impairment and resting-state network connectivity in Parkinson’s disease: Connectivity in Parkinson’s Disease,” Hum. Brain Mapp., vol. 36, no. 1, pp. 199–212, Jan. 2015, doi: 10.1002/hbm.22622.

[63] E. Cesnaite et al., “Alterations in rhythmic and non-rhythmic resting-state EEG activity and their link to cognition in older age,” NeuroImage, vol. 268, p. 119810, Mar. 2023, doi: 10.1016/j.neuroimage.2022.119810.

[64] K. Akbaş, K. Petrillo, H. Ehsani, and N. Toosizadeh, “Assessing aging-related changes of the brain–heart interconnection using functional near-infrared spectroscopy,” Clin. Neurophysiol., vol. 188, p. 2111925, Aug. 2026, doi: 10.1016/j.clinph.2026.2111925.

[65] S. Moguilner et al., “Brain clocks capture diversity and disparities in aging and dementia across geographically diverse populations,” Nat. Med., vol. 30, no. 12, pp. 3646–3657, Dec. 2024, doi: 10.1038/s41591-024-03209-x.

[66] S. Hoops et al., “Validity of the MoCA and MMSE in the detection of MCI and dementia in Parkinson disease”.

[67] M. Iniguez et al., “Heart-brain synchronization breakdown in Parkinson’s disease,” Npj Park. Dis., vol. 8, no. 1, p. 64, May 2022, doi: 10.1038/s41531-022-00323-w.

[68] J. L. Hazelton et al., “Thinking versus feeling: How interoception and cognition influence emotion recognition in behavioural-variant frontotemporal dementia, Alzheimer’s disease, and Parkinson’s disease,” Cortex, vol. 163, pp. 66–79, Jun. 2023, doi: 10.1016/j.cortex.2023.02.009.

[69] L. Ricciardi et al., “Know thyself: Exploring interoceptive sensitivity in Parkinson’s disease,” J. Neurol. Sci., vol. 364, pp. 110–115, May 2016, doi: 10.1016/j.jns.2016.03.019.

[70] P. C. Salamone et al., “Interoception Primes Emotional Processing: Multimodal Evidence from Neurodegeneration,” J. Neurosci., vol. 41, no. 19, pp. 4276–4292, May 2021, doi: 10.1523/JNEUROSCI.2578-20.2021.

[71] G. Santangelo et al., “Interoceptive processing deficit: A behavioral marker for subtyping Parkinson’s disease,” Parkinsonism Relat. Disord., vol. 53, pp. 64–69, Aug. 2018, doi: 10.1016/j.parkreldis.2018.05.001.

